# Prevalence and factors associated with advanced HIV disease among young people aged 15 - 24 years in a national referral hospital in Sierra Leone: A cross-sectional study

**DOI:** 10.1101/2023.11.07.23296880

**Authors:** Mamadu Baldeh, Samuel Kizito, Sulaiman Lakoh, Daniel Sesay, Frida Dennis, Umu Barrie, Samuel Adeyemi Williams, Dimbintsoa Rakotomalala Robinson, Franck Lamontagne, Franck Amahowe, Patrick Turay, Ozge Sensory-Bahar, Elvin Geng, Fred M. Ssewamala

## Abstract

**Background:** Advanced HIV in young people living with HIV is an increasingly pressing public health issue in sub-Saharan Africa. Despite global progress in early HIV testing and reducing HIV-related deaths, many young people with HIV continue to experience HIV disease progression in sub-Saharan Africa. This study provides an overview of the prevalence, clinical manifestations, and factors associated with advanced HIV in young people seeking medical services in a major hospital in Sierra Leone.

**Methods:** We used a cross-sectional design to collect data from HIV patients aged 15 to 24 years at a major hospital in Sierra Leone between September 2022 and March 2023. Advanced HIV was defined as (i) CD4+ below 200 cells/mm3 or (ii) WHO clinical stage 3 or 4. Logistic regression models determined the association between observable independent characteristics and advanced HIV. The statistical significance level was set at 0.05 for all statistical tests.

**Results:** About 40% (231/574) of patients were recruited; 70.6% (163/231) were inpatients, and 29.4% (68/231) were outpatients. The mean age was approximately 21.6 years (SD ±2.43). The overall prevalence of advanced HIV was 42.9% (99/231), 51.5% (35/68) of outpatients, and 39.3% (64/163) of inpatients. Age of inpatients (OR, 1.23; 95% CI, 1.00-1.52; p= 0.047) was associated with a higher risk. Female sex (OR, 0.51; 95% CI, 0.28-0.94; p= 0.030), higher education (OR, 0.27; 95% CI, 0.10 – 0.78; p= 0.015), and Body Mass (OR, 0.10; 95% CI, 0.01-0.77; p= 0.028) were at lower risk of advance HIV. Common conditions diagnosed in this population are tuberculosis (13.58%), hepatitis B (6.13%), Kaposi sarcoma (3.07%), and esophageal candidiasis (2.45%).

**Conclusion:** We reported a high prevalence of advanced HIV among young patients in a referral Hospital in Sierra Leone. This emphasises the need to strengthen public health measures and policies that address challenges of access to HIV services.

**Strengths and limitations of this study:**

- This is the first study in Sierra Leone and the sub-region to examine the burden of advanced HIV and its predictors in young people living with HIV.
- Calls for a more targeted approach to addressing gaps in health service delivery for young people living with HIV in Sierra Leone.
- The study is limited by the unavailability of several laboratory investigations to monitor patient progress.

## Introduction

Human immunodeficiency virus (HIV) remains a major global health challenge, affecting millions of people worldwide. In 2022, an estimated 39 million people were living with HIV (PLHIV) globally, with sub-Saharan Africa (SSA) carrying over 75% of the global disease burden (1,2). Despite the significant progress in scaling up HIV prevention, increased access to antiretroviral therapy (ART), and improved life expectancy of PLHIV, a systematic review of about 2,500 records showed an AHD prevalence of 43.5% among ART-naïve and 58.6% among ART-experienced (3). AHD, defined by WHO as PLHIV having a CD4 < 200 cells/mm^3^ or stage 3 or 4 for adults, adolescents, and older children, is associated with a high risk of death and increased healthcare expenditure (4).

In SSA, complex, multi-layered social dynamics and programmatic factors have been linked to the increased burden of AHD (5). Despite late-stage diagnosis, issues related to linkage to care, treatment interruptions, and social challenges such as stigma and discrimination contribute to the disease progression (6–8) and the occurrence of opportunistic infections(8,9).

Although significant progress in the past decade has resulted in a 46% decrease in new HIV infections among youths aged 15-24 years, nearly 3 million young people present late to healthcare facilities for care. AHD remains a public health concern in this population group. Indeed, adolescents with AHD present a greater risk of mortality from causes related to HIV compared to adults (6,7,10).

In Sierra Leone, the national HIV prevalence is estimated at 1.7% among people aged 15-49 (11,12). Although the prevalence has remained almost constant in the last 5 years, substantial efforts remain to be made to achieve the 95-95-95 goal, with only 76% of PLHIV in Sierra Leone knowing their HIV status and receiving antiretroviral therapy (ART) (13). The advent of the 10-year civil war, the 2014- 2016 Ebola epidemic, and other public health system challenges have made young people more vulnerable and, thus, have prompted the increased call to action to address the growing HIV epidemic nationally (14,15).

Much of the population is relatively young, with the individual median age of 19.9 years old (16,17)and roughly one-fourth of the entire population falling within the age bracket of 15 to 24 years (18). The impact of AHD on the well-being and socioeconomic status of YLHIV in Sierra Leone is yet to be fully understood. Despite these deep-rooted public health challenges, a PubMed search in June 2023 found no data on the burden of AHD among young people aged 15 - 24 years living with HIV in Sierra Leone. Studying the level of AHD burden and identifying contributing factors in YLHIV in Sierra Leone will aid global efforts to combat the HIV epidemic by developing well-informed policies and improving clinical practices. In this observational study, we aim to explore the prevalence of AHD and associated risk factors of AHD amongst young people aged 15 -24 years living with HIV at a national referral hospital in Sierra Leone.

## Methods

### Study Design and Study Setting

We used cross-sectional data from patients’ records living with HIV aged 15 to 24 at a tertiary hospital called Connaught Hospital, located in Freetown, Sierra Leone. Connaught Hospital is part of the University of Sierra Leone Teaching Hospitals Complex. It provides medical services to communities in Western Sierra Leone, including over 5000 outpatients and 14.8% of all inpatients admitted to this hospital (19). Given its status as the primary referral center in the country and its role as the main provider of advanced HIV care in Sierra Leone, Connaught Hospital is considered the largest HIV center in the country.

### Participant selection

Between September 2022 and March 2023, we collected inpatient and outpatient data on the medical wards and the HIV clinic. Patients were eligible to participate in the study if they met the following criteria: 1) aged 15-24 years, 2) diagnosed with HIV and status confirmed, and 3) receiving care at the study hospital during the study period as inpatients or outpatients. Inpatients were admitted to the medical wards after 24 hours of medical observation for further treatment, and outpatients presented to the HIV clinic for routine medical visits and care within 24 hours of presentation. Overall, 231/574 patients’ medical records were abstracted, including 163/163 inpatients and 68/411 outpatients. All inpatient participants met inclusion criteria and hence were included in the study.

### Measures and definitions

The study gathered data using a paper-based data extraction form and an electronic database established at the HIV clinic and medical wards by trained staff members. The collected data included demographic information, HIV details (Status, date of first diagnosis, medications, and investigations), and socio-economic characteristics. AHD was defined as any YLHIV with (i) CD4+ below 200 cells/mm^3^ or (ii) WHO clinical stage 3 or 4. We used patients’ clinical features to confirm opportunistic infections such as esophageal candidiasis and molluscum contagiosum.

We determined HIV status using a third-generation rapid diagnostic test (Standard Diagnostics Bioline HIV -1/2 3.0) and CD4 cell count using the Alere Pima Analyzer. Kaposi sarcoma was diagnosed histologically through the Kaposi sarcoma program in Sierra Leone. We made a diagnosis of tuberculosis (TB) using Xpert MTB Rif assay and/or urinary TB lipoarabinomannan.

### Data management and analysis

Data was inputted into a Microsoft Excel spreadsheet. All statistical analyses were performed using Stata version 17.0. Descriptive statistics were computed to summarize the data. Continuous variables were summarized using mean (± standard deviation) or median (interquartile range) according to their distribution, while categorical variables were presented as frequencies and proportions. We stratified the sample into two categories: those with AHD and those without AHD. The two groups were compared using independent samples t-tests for continuous variables and chi-squared tests for categorical variables.

To identify the potential correlates of AHD, we fitted two separate logistic regression models. The first model included the entire sample, encompassing both inpatients and outpatients. In contrast, the second model was fitted exclusively for the sub-sample of inpatients. In the logistic regression models, AHD served as the dependent variable, which was dichotomized, with the presence of AHD coded as 1 and absence coded as 0.

The logistic regression assumptions were thoroughly checked, including linearity in the logit for continuous independent variables using the Box-Tidwell procedure, where each continuous variable is paired with its natural log term and the absence of multicollinearity. The goodness-of-fit of our logistic regression models was assessed using the Hosmer-Lemeshow test. In this test, a non-significant result indicates a good fit of the model, as it suggests no difference between the observed and predicted values of the dependent variable in our model. Given the considerable correlation between substance use and smoking, we included an interaction term for these two variables in our model. This enabled us to investigate whether the effect of substance use on AHD differed depending on whether the participant was a smoker. The interaction term was created by multiplying the substance use and smoking variables and then included in our logistic regression models. The statistical significance level was set at an alpha of 0.05 for all tests. We reported the odds ratios and the 95% confidence intervals.

### Ethical consideration

Ethics approval was obtained from the Sierra Leone Ethics and Scientific Review Committee (SLESRC No. 032/09/2022). Data extraction was done from routine medical reports and did not require assenting or consenting—the data was fully de-identified and anonymized upon entry.

## Results

### Sociodemographic characteristics of the participants

Over the course of the study, 574 patients had a confirmed HIV status, 71.6% (411/574) HIV patients were seen in the HIV clinic (outpatients), and of these, 16.6% (68/411) were YLHIV. The remaining 28.4% (163/574) of the patients were inpatients living with HIV (Fig. 1).

**Figure 1:**
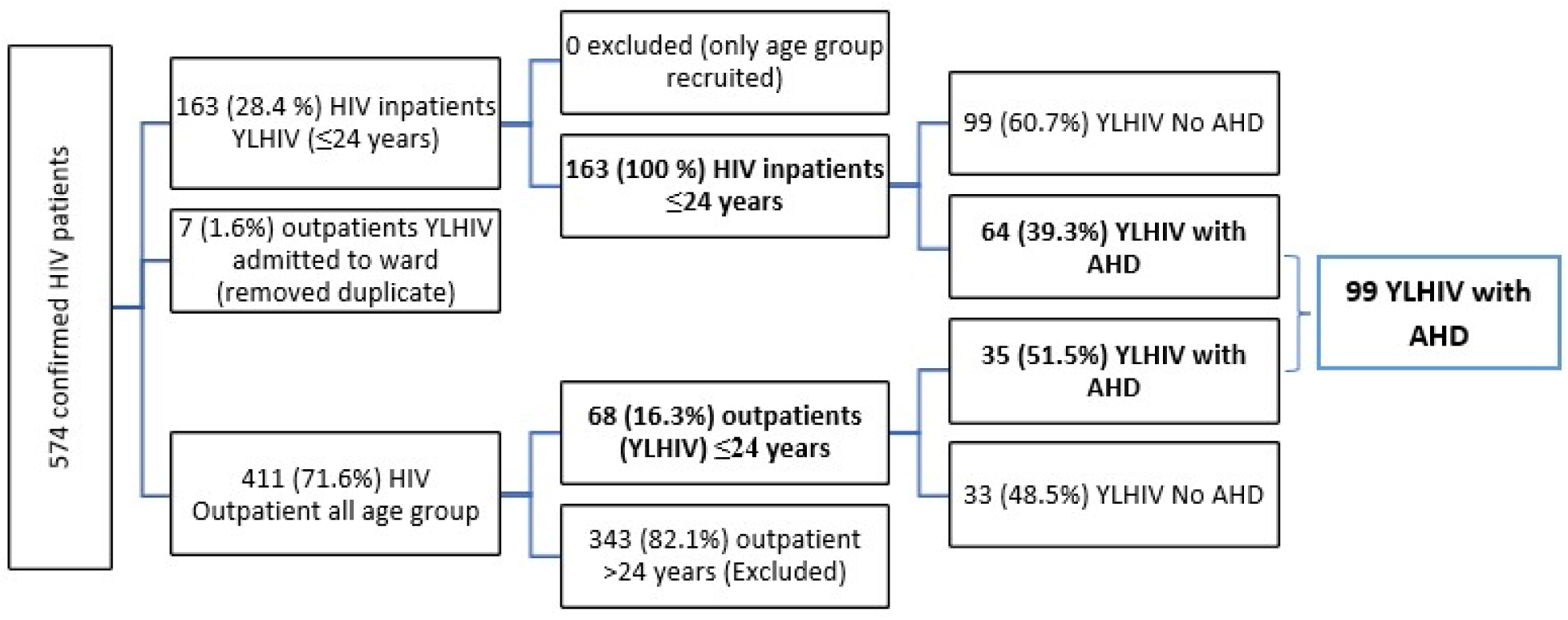
General characteristics of PLHIV seen at Connaught Hospital

**Figure 2:**
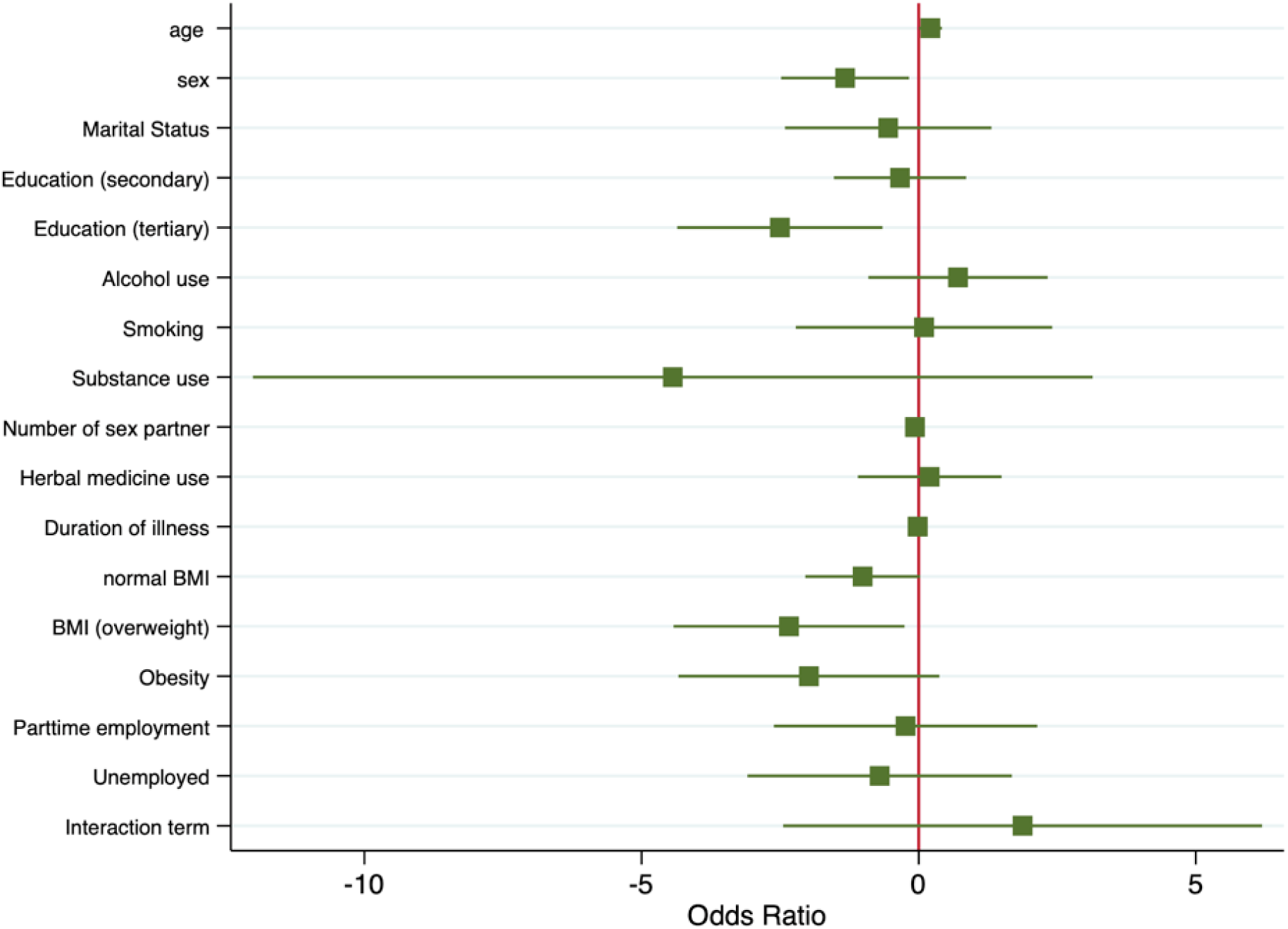
Forest plot for the logistic regression model assessing the association between various predictors and AHD among inpatient YLHIV

About 66.3% (108/163) of inpatients and 53.3% (43/75) of outpatients were female. The overall mean age for inpatients was 21.8 years (SD ±2.4), and for outpatients, 21.4 years (SD± 2.1). Most had at least secondary level education (60.9%), and more than 50% were unemployed with no source of constant income. About 20% used recreational substances. Over 95% of all patients were referred to Connaught Hospital from another public health facility, with 90% of all patients being classified as known HIV cases. HIV infection has been diagnosed for 5.2 months (SD ±7.3), and ART use was reported in approximately 80% of cases (Table 1 & 2).

**Table 1:**
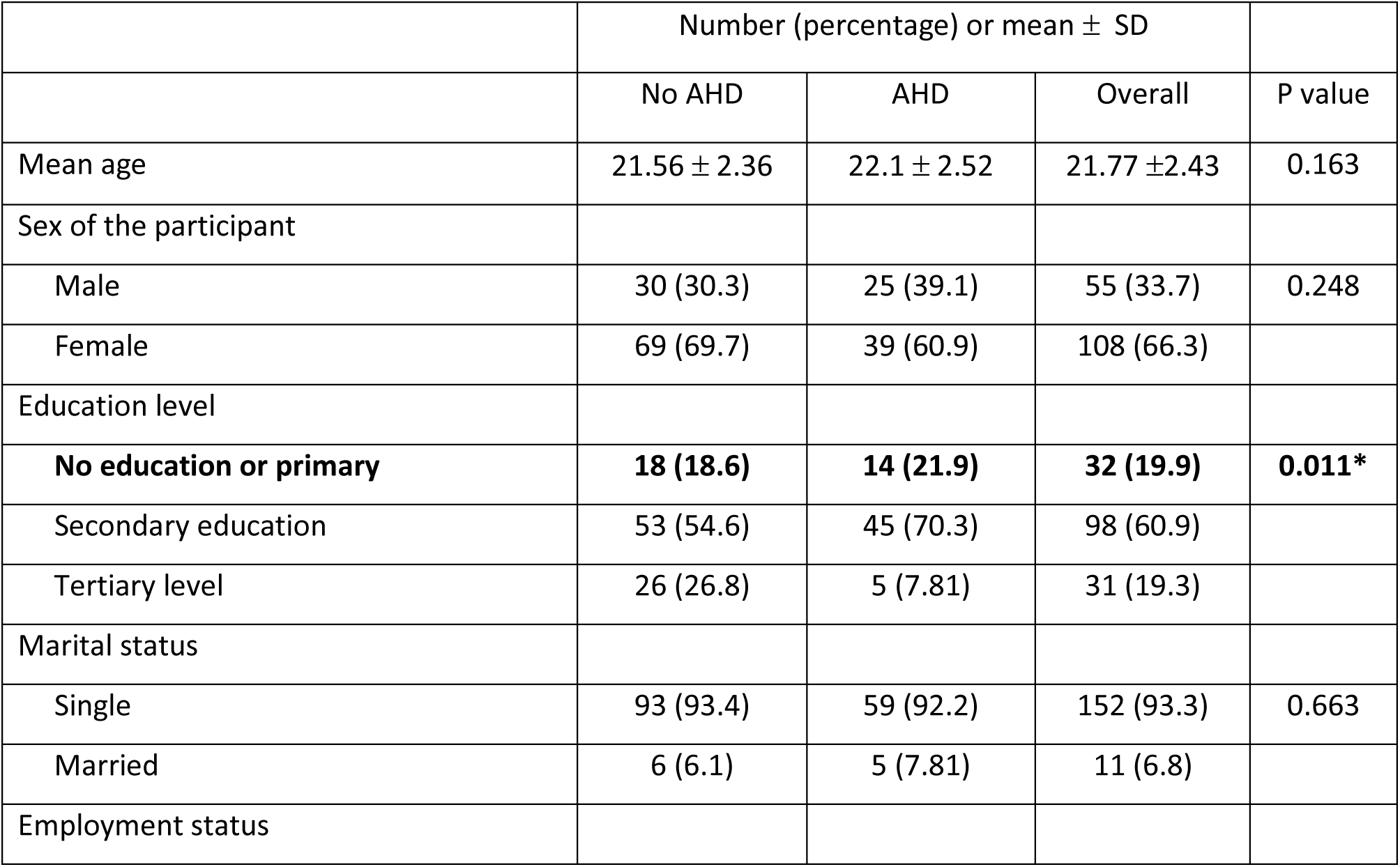
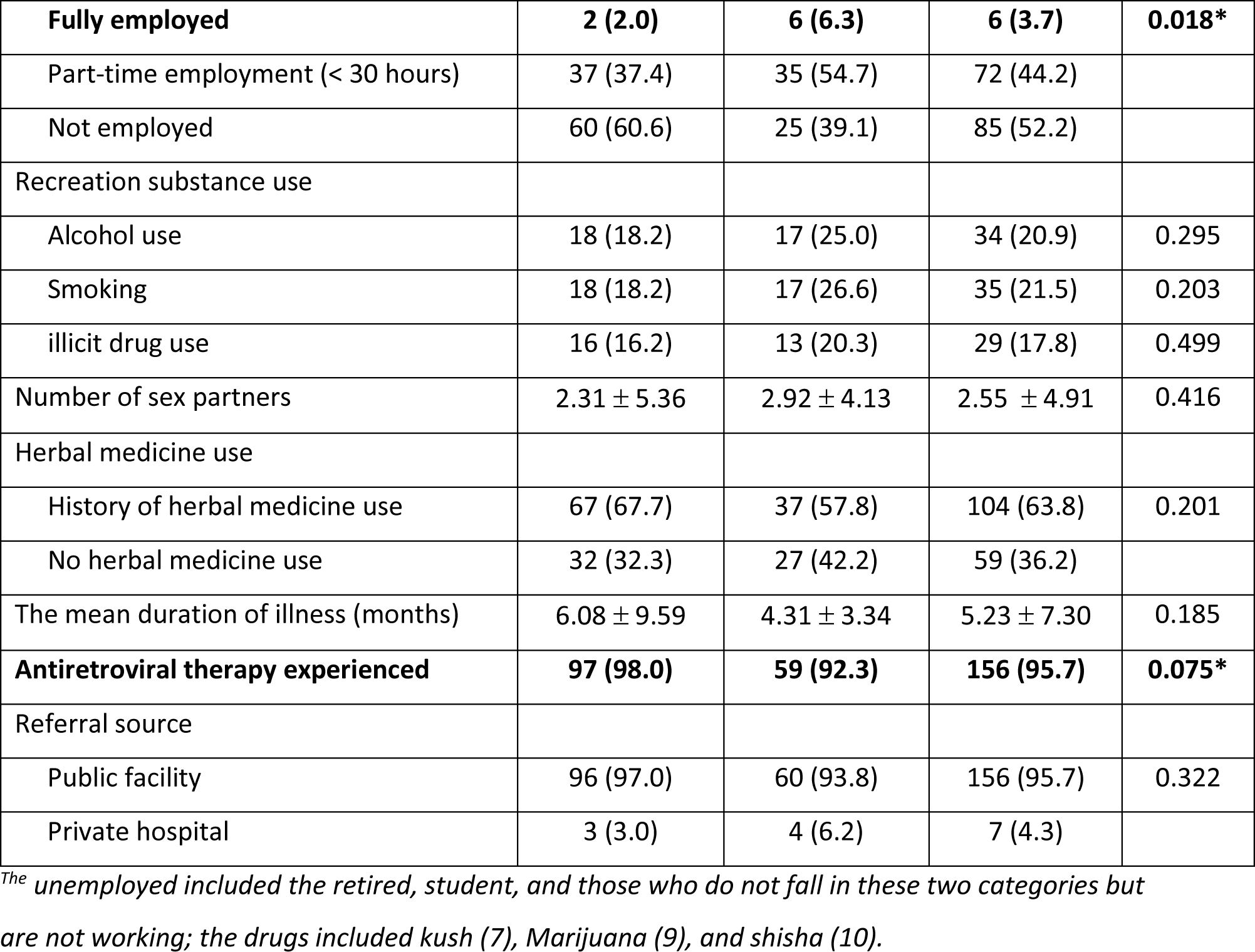
Sociodemographic characteristics of 163 inpatients YLHIV at Connaught Hospital.

**Table 2:** Sociodemographic characteristics of 68 outpatients YLHIV seeking HIV care services at Connaught Hospital.

|  | Number (percentage) or mean ± SD |  |  |  |
| --- | --- | --- | --- | --- |
|  | No AHD (n=33) | AHD (n=35) | Overall | P value |
| Mean age | 21.67 ±1.78 | 21.19 ±2.48 | 21.44 ±2.14 | 0.351 |
| Sex of the participant |  |  |  |  |
| Male | 13 (39.4) | 19 (54.3) | 32 (47.1) | 0.089 |
| Female | 20 (60.6) | 16 (45.7) | 36 (52.9) |  |
| Education level |  |  |  |  |
| No education or primary | 5 (15.1) | 9 (36.0) | 14 (20.6) | 0.317 |
| Secondary education | 23 (69.7) | 19 (54.3) | 42 (61.8) |  |
| Tertiary level | 9 (27.3) | 3 (8.6) | 12 (17.6) |  |
| Marital status |  |  |  |  |
| Single | 32 (97.0) | 33 (94.4) | 65 (95.6) | 0.509 |
| Married | 1 (3.0) | 2 (5.56) | 3 (4.4) |  |
| Employment status |  |  |  |  |
| Employed | 13 (39.4) | 14 (40.0) | 27 (39.7) | 0.115 |
| Not employed<sup>a</sup> | 20 (60.6) | 21 (60.0) | 41 (60.3) |  |
| Recreation substance use |  |  |  |  |
| Alcohol use | 10 (25.6) | 9 (25.0) | 19 (25.3) | 0.949 |
| Smoking | 6 (15.4) | 7 (19.4) | 13 (17.3) | 0.642 |
| illicit drug use<sup>b</sup> | 5 (12.8) | 1 (2.78) | 6 (8.00) | 0.109 |
| Antiretroviral therapy experienced | 32 (82.1) | 30 (83.3) | 62 (82.7) | 0.883 |

<sup>a</sup>The unemployed included the retired, student, and those who do not fall in these two categories but are not working; the drugs included kush (7), Marijuana (9), and shisha (10).

### AHD Prevalence

The overall prevalence of AHD in YLHIV was 42.9% (99/231); 51.5% (35/68) of outpatients, while 39.3% (64/163) of inpatients had AHD (Fig. I).

### Clinical presentations and co-morbidity among inpatients

We further explored the most common symptoms presented among inpatients (163). Weight loss (44.2%), fever (30.7%), cough (18.4%), headache (17.8%), and diarrhea (8.0%) were the most common symptoms, while TB (13.6%), chronic hepatitis B (6.1%), Kaposi sarcoma (3.1%), and esophageal candidiasis (2.5%) were the most common diagnosed co-infections or opportunistic diseases. Approximately 34% of patients had CD4+ cell count below 200 mm^3^, and 30% had a WHO clinical stage 3 or stage 4 disease (Table 3).

**Table 3:** Clinical characteristics of 163 youths living with HIV admitted to Connaught Hospital in Sierra Leone.

| Participant characteristics | Frequency (percentage) |
| --- | --- |
| Common presenting symptoms |  |
| Weight loss | 72 (44.2) |
| Fever | 50 (30.7) |
| Cough | 30 (18.4) |
| Headache | 29 (17.8) |
| Diarrhea | 13 (7.9) |
| Others<sup>a</sup> | 71 (43.6) |
| Body mass index |  |
| Underweight | 44 (27.2) |
| Normal | 98 (60.5) |
| Overweight | 12 (7.4) |
| Obesity | 8 (4.9) |
| Admission diagnosis |  |
| Tuberculosis | 22 (13.5) |
| Hepatitis B | 10 (6.1) |
| Kaposi Sarcoma | 5 (3.1) |
| Esophageal candidiasis | 4 (2.5) |
| Others<sup>b</sup> | 15 (9.2) |
| CD4 count |  |
| CD4+ below 200 | 41 (34.2) |
| CD4+ more than 200 | 79 (65.8) |
| WHO HIV clinical stage |  |
| Stage 1 | 66 (40.5) |
| Stage 2 | 47 (28.8) |
| Stage 3 | 43 (26.4) |
| Stage 4 | 7 (4.3) |
| Advanced HIV disease |  |
| No advanced HIV | 99 (60.7) |
| Diagnosed with advanced HIV | 64 (39.3) |

<sup>a</sup>Other symptoms include abdominal pain (16), genital discharge (14), difficult breathing (8), night sweats (7), genital ulcer (4), vomiting (3); <sup>b</sup>Other admission diagnoses include gastric ulcers, herpes labialis, molluscum contagiosum, PPE, post-herpetic neuralgia, urticaria, vaginal ulcer syndrome.

### Factors associated with AHD

Univariate analysis showed that the education level, employment status, and duration of Antiretroviral therapy were associated with AHD (p < 0.05) among inpatients.

Multivariate logistic regression analysis showed that the age of inpatients (OR, 1.23; 95% CI, 1.00-1.52; p= 0.047) was at higher risk of AHD than the age of all participants. Additionally, female sex (OR, 0.51; 95% CI, 0.28-0.94; p = 0.030), higher education (OR, 0.27; 95% CI, 0.10 – 0.78; p = 0.015), and higher body mass index (BMI) (OR, 0.10; 95% CI, 0.01-0.77; p= 0.028) were at a significantly lower risk of AHD in both groups (Table 4).

**Table 4:** Factors associated with advanced HIV disease among youths (15 – 24 years) living with HIV admitted to Connaught Hospital, Sierra Leone.

|  | Entire sample (n=231) |  |  | Inpatients (n=163) |  |  |
| --- | --- | --- | --- | --- | --- | --- |
| Characteristics | <u>Odds Ratio</u> | <u>95% CI</u> | <u>P value</u> | <u>Odds Ratio</u> | <u>95% CI</u> | <u>P value</u> |
| Participant age | <u>1.03</u> | <u>0.91 – 1.17</u> | <u>0.611</u> | <u>1.23</u> | <u>1.00 – 1.52</u> | <u>0.047 *</u> |
| Sex of the participant |  |  |  |  |  |  |
| <u>Male</u> | <u>1</u> |  |  | <u>1</u> |  |  |
| <u>Female</u> | <u>0.51</u> | <u>0.28 – 0.94</u> | <u>0.030 *</u> | <u>0.27</u> | <u>0.08 – 0.84</u> | <u>0.024 *</u> |
| Marital status |  |  |  |  |  |  |
| <u>Single</u> | <u>1</u> |  |  | <u>1</u> |  |  |
| <u>Married</u> | <u>1.24</u> | <u>0.37 – 4.16</u> | <u>0.730</u> | <u>0.58</u> | <u>0.09 – 3.71</u> | <u>0.562</u> |
| Education level |  |  |  |  |  |  |
| <u>No education or primary</u> | <u>1</u> |  |  | <u>1</u> |  |  |
| <u>Secondary education</u> | <u>0.85</u> | <u>0.41 – 1.76</u> | <u>0.667</u> | <u>0.71</u> | <u>0.22 – 2.36</u> | <u>0.580</u> |
| <u>Tertiary level</u> | <u>0.27</u> | <u>0.10 – 0.78</u> | <u>0.015 *</u> | <u>0.08</u> | <u>0.01 – 0.52</u> | <u>0.008 *</u> |
| Recreation substance use |  |  |  |  |  |  |
| <u>Alcohol use</u> | <u>1.31</u> | <u>0.56 – 3.09</u> | <u>0.533</u> | <u>2.03</u> | <u>0.40 – 10.23</u> | <u>0.390</u> |
| <u>Smoking</u> | <u>2.56</u> | <u>0.62 – 11.6</u> | <u>0.195</u> | <u>1.10</u> | <u>0.11 – 11.14</u> | <u>0.935</u> |
| <u>illicit drug use</u> | <u>0.60</u> | <u>0.01 – 117.0</u> | <u>0.851</u> | <u>0.01</u> | <u><0.01 – 23.07</u> | <u>0.251</u> |
| Number of sex partners |  |  |  | <u>0.94</u> | <u>0.86 – 1.01</u> | <u>0.104</u> |
| Herbal medicine use |  |  |  |  |  |  |
| <u>No herbal medicine use</u> |  |  |  | <u>1</u> |  |  |
| <u>History of herbal medicine</u> |  |  |  | <u>1.22</u> | <u>0.33 – 4.46</u> | <u>0.764</u> |
| Duration of illness |  |  |  | <u>0.98</u> | <u>0.91 – 1.06</u> | <u>0.669</u> |
| Body mass index |  |  |  |  |  |  |
| <u>Underweight</u> |  |  |  | <u>1</u> |  |  |
| <u>Normal</u> |  |  |  | <u>0.36</u> | <u>0.13 – 1.02</u> | <u>0.056</u> |
| <u>Overweight</u> |  |  |  | <u>0.10</u> | <u>0.01 – 0.77</u> | <u>0.028 *</u> |
| <u>Obesity</u> |  |  |  | <u>0.14</u> | <u>0.01 – 1.45</u> | <u>0.099</u> |
| <u>Employment status</u> |  |  |  |  |  |  |
| <u>Fully employed</u> | <u>1</u> |  |  | <u>1</u> |  |  |
| <u>Part time employment</u> | <u>0.36</u> | <u>0.06 – 2.28</u> | <u>0.277</u> | <u>0.78</u> | <u>0.07 – 8.50</u> | <u>0.845</u> |
| <u>Not employed</u> | <u>0.30</u> | <u>0.05 – 1.95</u> | <u>0.205</u> | <u>0.49</u> | <u>0.05 – 5.38</u> | <u>0.563</u> |
| <u>Substance*smoking interaction</u> | <u>0.66</u> | <u>0.04 – 12.36</u> | <u>0.783</u> | <u>6.51</u> | <u>0.09 – 4.89.34</u> | <u>0.395</u> |
|  | <u>Hosmer Lemeshow Goodness of fit:</u><br><u>$\chi^2 (100) = 122.81, p = 0.061.$</u> |  |  | <u>Hosmer Lemeshow Goodness of fit:</u><br><u>$\chi^2 (96) = 109.35, p = 0.166.$</u> |  |  |

*\* Denotes associations that are statistically significant*

## Discussions

This first study in Sierra Leone and the West African sub-region to assess the prevalence and risk factors of AHD in YLHIV reported an AHD prevalence of 42.9% (36.6% - 49.4%) among 231 YLHIV between September 2022 and February 2023 at the national referral hospital in Sierra Leone. Female sex, higher educational level, age of inpatients, and higher BMI of inpatients were independent predictors of AHD.

We could not directly compare our findings to other AHD studies because there are limited AHD studies in young adults. However, despite the low national prevalence of HIV of 1.7% in Sierra Leone, the prevalence of AHD among young people in this study was higher than that reported in many other countries in sub-Saharan Africa. A study in South Africa, Malawi, and Kenya reported a low AHD prevalence of 9.7% (20,21) Another study in Kenya reported an AHD prevalence of 33% (20). The higher prevalence of AHD in young people in this study reflects the operational realities of the health sector response to HIV services in Sierra Leone. It contextualizes warnings from the International AIDS Society that countries in the West and Central African regions lag behind others in meeting the 95– 95-95 global target (22,23) Therefore, the HIV program should redouble its efforts to address known barriers, such as the low HIV testing rate, low rates of HIV viral suppression, the high burden of opportunistic fungal and non-fungal infections, and stroke (24–28).

More outpatients than inpatients in this study have AHD (49.4% vs. 36.6%). This finding underscores the fact that CD4+ count and screening for opportunistic infections such as TB and cryptococcal disease are recommended in AHD patients regardless of the setting in which they are assessed (29) However, the declining trend in CD4 testing has been observed across sub-Saharan Africa. This trend poses a risk of overlooking AHD diagnoses, potentially jeopardizing the effectiveness of antiretroviral therapy (ART) programs (29).

We found that age was an independent predictor of AHD among inpatients. Multiple studies (30–32) have highlighted the effect of age on AHD and emphasize disease progression not only in the elderly but even in young people who could have been infected for quite a long period in Sierra Leone’s context, facing multiple socio-economic and health service challenges. This fact highlights the need to include young people among HIV testing targets and treatment to prevent HIV disease progression among young HIV patients in healthcare settings. The level of education also shows its association with disease progression. This could be attributed to seeking medical services and a better understanding of instructions, including medication intervals and clinical follow-up visits, which have also been reported in multiple studies (33–35). Our study is also consistent with findings from other studies that show that more women are less likely than men to present with AHD (36,37)

TB is the most common co-infection in inpatients with AHD in this study, reflecting the high burden of TB in this hospital (38) TB is a common opportunistic infection in PLHIV, and the high TB prevalence among YLHIV is not unique to our setting as it has also been documented in other low-and middle-income countries (39,40). The hepatitis B co-infection rate of 6% in YLHIV reported in this study is expected and could be explained by the fact that hepatitis and HIV share common transmission routes. We are concerned about this finding because co-infection with TB and hepatitis B in YLHIV can significantly affect the progression to AHD and the development of comorbidities such as immune reconstitution disease and hepatocellular carcinoma (39,40) Furthermore, a recent study (24) in Sierra Leone showed a high prevalence of stroke in HIV patients with lower CD4 cell counts. Similarly, another study observed that HIV co-infection with TB and hepatitis B co-infection are strongly associated with prediabetes, diabetes, and liver fibrosis/cirrhosis (41). Hence, we should use these facts as a unique opportunity to scale up hepatitis B prevention programs and for the integration of comprehensive care of comorbidities in HIV service delivery points in Sierra Leone.

Our study has strengths and limitations. First, this study was conducted in one hospital, and its findings cannot be generalized to the YLHIV population in Sierra Leone. Second, we are limited by the unavailability of several laboratory investigations to monitor patient progress. Nonetheless, this is the first study in Sierra Leone and the West African sub-region to examine the burden of AHD and its predictors in YLHIV, calling for a more targeted approach to addressing gaps in health service delivery for YLHIV in Sierra Leone.

## Conclusion

We report a high prevalence of AHD among YLHIV in the national referral hospital in Sierra Leone. These findings highlight the need to strengthen public health measures and policies to address socioeconomic barriers limiting access to healthcare services, particularly for YLHIV. Through concerted efforts, we call for the implementation of measures to improve access to timely diagnosis and better access to treatment among young people.

## Authors contributions

MB and FS conceptualized and designed the research study. MB, UB, DS, and FD collected the data required for the study. MB, SK, and SL were responsible for drafting the initial version of the manuscript, conducting data analysis, and interpreting the results. All authors have critically reviewed multiple manuscript versions, provided significant edits and feedback, and approved the final version for submission. FS, as the Principal Investigator (PI) and mentor, oversaw the entire ACHIEVE project, providing a critical review, guidance, and leadership throughout the research process.

## Data Availability

All data produced in the present study are available upon reasonable request to the authors.

## Acknowledgment

We thank all ICHAD (International Center for Child Health and Development) members at Brown School at Washington University in St. Louis and program managers for ACHIEVE program, Laura Peer, Chelsea Hand-Sheridan, Bethel Mandefro, Portia Nartey, Rebecca Esliker, University of Makeni, Geng Lab and Connaught Hospital.

